# Hemodilution in High Risk Cardiac Surgery: Laboratory Values, Physiological Parameters and Outcomes

**DOI:** 10.1101/2021.07.14.21260529

**Authors:** Domagoj Mladinov, Luz A Padilla, Benjamin Leahy, Joseph B Norman, Jacob Enslin, Riley S Camp, Kyle W Eudailey, Kenichi Tanaka, James E Davies

## Abstract

**Background:** Acute normovolemic hemodilution (ANH) is a blood conservation strategy in cardiac surgery, predominantly used in coronary artery bypass graft (CABG) and/or valve procedures. Although higher complexity cardiac procedures may benefit from ANH, concerns for hemodynamic instability and organ injury during hemodilution hinder its wider acceptance. Laboratory and physiological parameters during hemodilution in complex cardiac surgeries have not been described.

**Study Design and Methods:** This observational cohort (2019-2021) study included 169 patients who underwent thoracic aortic repair, multiple valve procedure, concomitant CABG with the aforementioned procedure, and/or redo sternotomies. Patients who received allogeneic blood were excluded. Statistical comparisons were performed between ANH (N=66) and non ANH controls (N=103). ANH consisted of removal of blood at the beginning of surgery and its return after cardiopulmonary bypass (CPB).

**Results:** Intraoperatively, the ANH group received more albumin (*p*=0.04) and vasopressor medications (*p*=0.01); while urine output was no different between ANH and controls. Bilateral cerebral oximetry (rSO_2_) values were similar before and after hemodilution. During bypass rSO_2_ were discretely lower in the ANH vs. control group (right rSO_2_ *p=*0.03, left rSO_2_ (*p*=0.05). No differences in lactic acid values were detected across the procedural continuum. Postoperatively, no differences in extubation times, ICU length of stay, kidney injury, stroke or infection were demonstrated.

**Discussion:** This study suggests hemodilution to be a safe and comparable blood conservation technique, even without accounting for potential benefits of reduced allogenic blood administration. The study may contribute to better understanding and wider acceptance of ANH protocols in high risk cardiac surgeries.

## INTRODUCTION

Acute normovolemic hemodilution (ANH) is a blood conservation technique utilized in cardiac surgery, which involves intraoperative removal of whole blood at the beginning of the procedure, volume replacement with crystalloid and/or colloid solutions, and return of the autologous blood after separation from cardiopulmonary bypass (CPB). ^1 2^ Although evidence demonstrating reduction in allogeneic blood transfusions with ANH is not definitive, a substantial number of retrospective studies and meta-analyses support its transfusion-sparing effect. ^3 4^ The large majority of those studies have been performed in coronary artery bypass graft (CABG) surgeries and single valve replacement/repairs, while results in more complex, higher risk cardiac surgeries are lacking. This may in part be attributed to increased concerns for hemodynamic instability and decreased oxygen delivery, secondary to blood removal and dilutional anemia in those procedures.^5 6^ However, some studies suggest that complex cardiac surgeries, such as aortic repairs, may particularly benefit from ANH, without increasing the risk of adverse outcomes. ^7 8^ As blood transfusions have been associated with increased morbidity and mortality, and allogeneic blood remains a limited resource, there is a need to consider expanding the practice of hemodilution to complex higher risk procedures. ^9 10^ This study was aimed to explore if ANH would not induce clinically significant changes in laboratory and physiological (hemodynamic) parameters in high-risk cardiac surgery. The results from this study can be useful to better understand and advance the acceptance of ANH protocols in high risk cardiac surgeries.

## MATERIALS AND METHODS

### Study Design and Patients

The study was approved by our Institutional Review Board (IRB-300005436). This was a single center observational cohort study that included 534 patients who underwent complex cardiac surgical procedures in the time period between 01/01/2019 and 01/15/2021 at a tertiary academic center. The following elective or emergent procedures were included: surgical repair of the thoracic aorta, multiple valve replacement/repair, concomitant single valve procedure and aortic repair and/or CABG, concomitant CABG and aortic repair and/or valve procedure; single valve- or CABG-only procedures were included only if they were repeat sternotomies.

The following exclusion criteria were used: patients who received allogeneic blood products intra-operatively or post-operatively during the index hospitalization, pre-operative hematocrit < 30%, known pre-existing coagulopathy or ongoing anticoagulant use, pre-operative left ventricular ejection fraction <35%, and pre-operative hemodynamic instability (requiring volume resuscitation, vasoactive medications and/or mechanical circulatory support). Through the whole study, surgical and perfusion techniques, as well as transfusion practices remained unchanged. Electronic medical records were reviewed and data for the study variables was manually extracted.

### Blood Management Practices

During the study period standard institutional blood conservation strategies included restrictive intra-operative fluid therapy, retrograde autologous priming of the CPB circuit, ultrafiltration during CPB, cell salvage and use of antifibrinolytic medications (tranexamic acid, total of 3 grams). Implementation of ANH was guided by the following institutional criteria: pre-operative hematocrit > 30%, no known pre-existing coagulopathy or ongoing anticoagulant use, preoperative left ventricular ejection fraction >35%, and hemodynamic stability.

Volume of collected autologous whole blood (AWB) was determined according to a nomogram based on patient’s ideal body weight and starting hematocrit. ^11^ Autologous blood was removed by gravity drainage via a large bore central venous catheter port, immediately after the catheter insertion. Blood was drained into citrate-phosphate-dextrose bags, and its weight continuously measured on an electronic scale during collection. Collection was stopped when the pre-determined amount of blood was obtained or when hemodynamic instability occurred. A maximum of 450 grams was allowed per bag. Standard hemodynamic monitors and transesophageal echocardiography (TEE) were used to guide fluid replacement (crystalloid and/or colloid solution, typically at 1:1 to 1:2 ratio) and if needed titration of vasopressor therapy. Autologous blood was kept in the operating room at room temperature, periodically gently rocked, and transfused back after protamine administration following separation from CPB.

### Laboratory Tests

Intra-operative laboratory tests included lactic acid, hemoglobin, hematocrit, platelet count, fibrinogen level, prothrombin time (PT), international normalized ration (INR), activated partial thromboplastin time (PTT). Post-operative hemoglobin, hematocrit, platelet count, creatinine (Cr), blood urea nitrogen (BUN) and calculated glomerular filtration rate (GFR) were collected.

### Physiological Parameters

Total volume of crystalloid and colloid solutions administered intra-operatively was recorded. Maximal infusion rate of vasopressor medications through the procedure were recorded and expressed as norepinephrine equivalents.^12 13^ Vasoactive medications included phenylephrine (Phe), vasopressin (Vaso), norepinephrine (NE) and epinephrine (Epi), and were titrated according to standard institutional practice. Regional cerebral oxygen saturation (rSO_2_) was continuously measured with near infrared spectroscopy (NIRS) monitors (INVOS™; Medtronic, Minneapolis, MN), and the highest intraoperative values during different stages of the procedure were determined. The NIRS monitor probes were placed on the right and left forehead, as recommended by the manufacturer. Urine output volume during the intra-operative period was also recorded.

### Postoperative Outcomes

The following post-operative outcomes were analyzed: chest tube output during the first 12h after surgery, re-operation for bleeding, intensive care unit (ICU) length of stay, duration of intubation, incidence of acute kidney injury (AKI), need for renal replacement therapy, ischemic stroke, and infection rate during the index hospitalization.

### Statistical Analysis

Patient demographics, peri-operative laboratory, physiological parameters and clinical outcomes were summarized (frequencies, percentages, medians, interquartile ranges [Q1-Q3], means, standard deviations) and compared using chi-square and Fisher’s exact for categorical variables, student t-test and Wilcoxon’s rank sum test for continuous variables where appropriate. Paired t-test was used to compare the laboratory values after separation from CPB among ANH patients only. Analysis of variance was used to compare the differences in mean peri-operative hospital laboratory results in both ANH and controls. Statistical Analytical Software (SAS) 9.4 (SAS institute, Cary, NC) was used for the analysis and all statistical tests of a two-sided *P* value of < 0.05 were considered significant.

## RESULTS

Between January 2019 and 2021, a total of 534 patients who underwent one of the high risk cardiac surgical procedures included in this study, of which 339 (63.5%) were treated with standard institutional blood conservations practices (control group), and 195 (36.5%) with ANH in addition to standard practices (ANH group). Inclusion criteria were met by a total of 169 patients: 103 (60.9%) in the control group and 66 (39.1%) in the ANH group. The average volume of removed AWB in the hemodilution group was 935 ± 253 ml.

There was no statistical difference in the basic demographic features of the two groups (Table 1). In the past medical history, the incidence of coronary artery disease was higher in the control compared to the ANH group (55.3% vs. 37.8% respectively; *p*=0.02). No difference between pre-operative laboratory tests, including kidney function tests, hemoglobin and coagulation studies, was detected. Finally, type of procedure and intra-operative procedural characteristics were not statistically different (Table 1).

**Table 1.** Demographics, Pre- and Intra- operative characteristics for hemodiluted patients and controls.

|  | Hemodiluted<br>N=66 (39.1%) | Controls<br>N=103 (60.9%) | *p* value |
| --- | --- | --- | --- |
| Age (years) | 58.3 ± 12.3 | 60.1 ± 14.8 | 0.41 |
| Male (%) | 51 (77.3) | 79 (77.4) | 0.97 |
| Weight (kg) | 93.6 ± 24.1 | 93.0 ± 21.4 | 0.86 |
| BMI (kg/m<sup>2</sup>) | 29.9 ± 6.8 | 30.6 ± 6.4 | 0.49 |
| Past medical history |  |  |  |
| CAD/MI | 25 (37.8) | 57 (55.3) | 0.02 |
| HTN | 54 (81.8) | 76 (73.8) | 0.22 |
| CVA | 10 (15.1) | 13 (12.6) | 0.63 |
| COPD | 11 (16.6) | 26 (25.2) | 0.18 |
| DM | 10 (15.1) | 29 (28.2) | 0.05 |
| CKD | 9 (13.6) | 11 (10.6) | 0.56 |
| Current tobacco use | 11 (16.7) | 17 (16.5) | 0.97 |
| EuroSCORE (%)* | 2.0 [1.3-3.4] | 2.2 [1.4-4.4] | 0.40 |
| ***Pre-Operative laboratory values*** |  |  |  |
| Creatinine (mg/dL) | 1.0 [0.9-1.3] | 1.0 [0.8-1.2] | 0.10 |
| BUN (mg/dL) | 18.6 ± 5.9 | 18.2 ± 7.8 | 0.76 |
| GFR < 60 mL/min | 19 (28.7) | 23 (22.3) | 0.34 |
| Hematocrit (%) | 41.3 ± 4.4 | 41.1 ± 4.6 | 0.81 |
| Hemoglobin (g/dL) | 13.5 ± 1.5 | 13.4 ± 1.5 | 0.53 |
| Platelet (10<sup>3</sup>/cmm) | 224.1 ± 63.5 | 215.7 ± 57.5 | 0.37 |
| INR | 1.02 [0.99-1.08] | 1.01 [0.96-1.05] | 0.27 |
| PT (seconds) | 13.4 ± 0.8 | 13.3 ± 0.7 | 0.15 |
| PTT (seconds) | 32.8 ± 13.2 | 31.1 ± 7.4 | 0.35 |
| ***Intra-Operative characteristics*** |  |  |  |
| Re-do sternotomy | 16 (24.2) | 24 (23.3) | 0.88 |
| Emergent | 2 (3.0) | 0 (0) | 0.15 |
| Dissection | 2 (3.0) | 0 (0) | 0.15 |
| Aneurysm | 33 (50.0) | 42 (40.7) | 0.23 |
| CABG | 23 (34.8) | 43 (41.7) | 0.36 |
| AVR | 38 (57.5) | 73 (70.8) | 0.07 |
| MVR | 12 (18.2) | 18 (17.6) | 0.92 |
| TVR | 3 (4.5) | 3 (2.9) | 0.67 |
| Blood removed (ml) | 935.2 ± 252.9 | - | - |
| Procedure time (min) | 193.0 ± 52.5 | 185.9 ± 53.6 | 0.39 |
| Cardiopulmonary bypass time (min) | 73.3 ± 29.4 | 65.2 ± 28.3 | 0.07 |
| Cross clamp time (min) | 54.9 ± 22.9 | 51.7 ± 20.7 | 0.35 |
| Circulatory arrest (yes) | 3 (4.5) | 1 (0.9) | 0.30 |

\*~50% of the data is missing; ^
Key: N (%); median [Q1-Q3]; mean ± SD; BMI=Body Mass Index, CAD=Coronary Artery Disease, MI=Myocardial Infarction, HTN=Hypertension, CVA=Cerebrovascular Accident, COPD=Chronic Obstructive Pulmonary Disease, DM=Diabetes Mellitus Type 2, CKD=Chronic Kidney Disease, BUN=Blood Urea Nitrogen, GFR=Glomerular Filtration Rate, INR=International Normalized Ratio, PT=Prothrombin time, PTT=Partial thromboplastin time, CABG=Coronary Artery Bypass Graft, AVR/MVR/TVR=Aortic/Mitral/Tricuspid Valve Replacement/Repair

After separation from CPB and protamine administration in the hemodiluted group, tests obtained before and after return of AWB demonstrated increase in mean hemoglobin (g/dL) from 9.2 ± 1.3 to 10.0 ± 1.4, hematocrit (%) 28.5 ± 4.1 to 30.8 ± 4.5, decreased median INR from 1.79 [1.60-1.90] to 1.56 [1.46-1.71], increased platelets (10^3/cmm) from 134.9 ± 48.5 to 149.4 ± 50.4, all with p=<.0001, while increase in fibrinogen and PTT were not significant (Table 2). Comparison of laboratory studies between the hemodiluted group after autologous blood return and the control group after separation from CPB showed no statistical difference in hematocrit, hemoglobin, INR, platelet count, PPT but did have a higher fibrinogen (mg/dL) in the latter group (229.9 ± 101.4 vs. 271.0 ± 106.3, p=0.01) (Table 2).

**Table 2.** Laboratory values after separation from cardiopulmonary bypass (CPB) in patients who were hemodiluted and controls

|  | Hemodiluted<br>N=66 (39.1%) |  |  | Controls<br>N=103 (60.9%) |  |
| --- | --- | --- | --- | --- | --- |
|  | Post CPB | Post ANH | *p* value* | Post CPB | *p* value** |
| Hematocrit (%) | 28.5 ± 4.1 | 30.8 ± 4.5 | <.0001 | 32.0 ± 4.3 | 0.10 |
| Hemoglobin (g/dL) | 9.2 ± 1.3 | 10.0 ± 1.4 | <.0001 | 10.4 ± 1.3 | 0.10 |
| Platelets (10<sup>3</sup>/cmm) | 134.9 ± 48.5 | 149.4 ± 50.4 | <.0001 | 148.6 ± 45.7 | 0.91 |
| Fibrinogen (mg/dL) | 223.6 ± 95.2 | 229.9 ± 101.4 | 0.31 | 271.0 ± 106.3 | 0.01 |
| INR | 1.79 [1.60-1.90] | 1.56 [1.46-1.71] | <.0001 | 1.56 [1.46-1.66] | 0.89 |
| PT (seconds) | 20.1 ± 1.8 | 18.9 ± 2.1 | <.0001 | 18.4 ± 1.5 | 0.07 |
| PTT (seconds) | 34.1 ± 4.3 | 38.6 ± 24.2 | 0.24 | 36.3 ± 24.2 | 0.56 |

Key: N (%); medians [Q1-Q3]; mean ± SD, INR=International Normalized Ratio, PT=Prothrombin time, PTT=Partial thromboplastin time, ANH=Acute Normovolemic Hemodilution
\*P value for Post ANH and Post CPB comparison among those who were hemodiluted only (n=66)
\*\*P value comparing the laboratory values Post ANH among those who were hemodiluted vs the Post CPB values among controls

Post-operatively, laboratory studies were analyzed at three different time points: upon arrival in the ICU, 8-36 hours after arrival to the ICU, and post-operative day 7 or last day of hospitalization (if discharged before 7 days). Laboratory values were comparable and had no statistical difference between the ANH and control groups in hemoglobin, hematocrit, platelet, INR, PTT, Cr, GFR or BUN, at any of the three post-operative time points (Table 3). Changes in hematocrit and hemoglobin concentration at multiple time points before, during and after surgery within the ANH and control groups were statistically different (*p*=<.0001, Figure 1). Similarly, multiple standard coagulation tests showed that within each group there was a significant change difference in platelet count, INR and PTT at various time points through the peri-operative period (Figure 2).

**Table 3.**
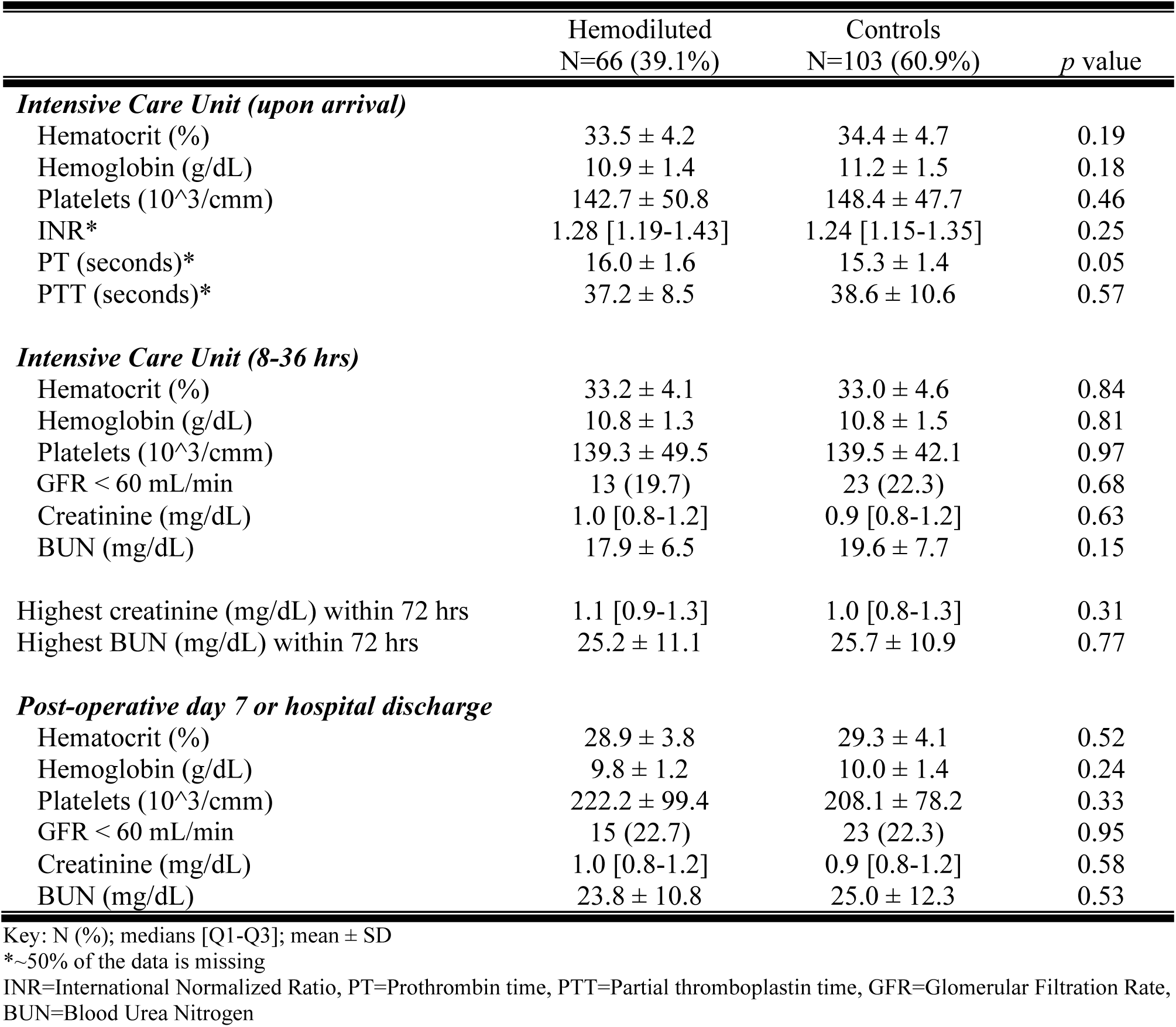
Post-operative laboratory values in patients who were hemodiluted and controls

|  | Hemodiluted<br>N=66 (39.1%) | Controls<br>N=103 (60.9%) | *p* value |
| --- | --- | --- | --- |
| ***Intensive Care Unit (upon arrival)*** |  |  |  |
| Hematocrit (%) | 33.5 ± 4.2 | 34.4 ± 4.7 | 0.19 |
| Hemoglobin (g/dL) | 10.9 ± 1.4 | 11.2 ± 1.5 | 0.18 |
| Platelets (10<sup>3</sup>/cmm) | 142.7 ± 50.8 | 148.4 ± 47.7 | 0.46 |
| INR* | 1.28 [1.19-1.43] | 1.24 [1.15-1.35] | 0.25 |
| PT (seconds)* | 16.0 ± 1.6 | 15.3 ± 1.4 | 0.05 |
| PTT (seconds)* | 37.2 ± 8.5 | 38.6 ± 10.6 | 0.57 |
| ***Intensive Care Unit (8-36 hrs)*** |  |  |  |
| Hematocrit (%) | 33.2 ± 4.1 | 33.0 ± 4.6 | 0.84 |
| Hemoglobin (g/dL) | 10.8 ± 1.3 | 10.8 ± 1.5 | 0.81 |
| Platelets (10<sup>3</sup>/cmm) | 139.3 ± 49.5 | 139.5 ± 42.1 | 0.97 |
| GFR < 60 mL/min | 13 (19.7) | 23 (22.3) | 0.68 |
| Creatinine (mg/dL) | 1.0 [0.8-1.2] | 0.9 [0.8-1.2] | 0.63 |
| BUN (mg/dL) | 17.9 ± 6.5 | 19.6 ± 7.7 | 0.15 |
| Highest creatinine (mg/dL) within 72 hrs | 1.1 [0.9-1.3] | 1.0 [0.8-1.3] | 0.31 |
| Highest BUN (mg/dL) within 72 hrs | 25.2 ± 11.1 | 25.7 ± 10.9 | 0.77 |
| ***Post-operative day 7 or hospital discharge*** |  |  |  |
| Hematocrit (%) | 28.9 ± 3.8 | 29.3 ± 4.1 | 0.52 |
| Hemoglobin (g/dL) | 9.8 ± 1.2 | 10.0 ± 1.4 | 0.24 |
| Platelets (10<sup>3</sup>/cmm) | 222.2 ± 99.4 | 208.1 ± 78.2 | 0.33 |
| GFR < 60 mL/min | 15 (22.7) | 23 (22.3) | 0.95 |
| Creatinine (mg/dL) | 1.0 [0.8-1.2] | 0.9 [0.8-1.2] | 0.58 |
| BUN (mg/dL) | 23.8 ± 10.8 | 25.0 ± 12.3 | 0.53 |

Key: N (%); medians [Q1-Q3]; mean ± SD
\*~50% of the data is missing
INR=International Normalized Ratio, PT=Prothrombin time, PTT=Partial thromboplastin time, GFR=Glomerular Filtration Rate, BUN=Blood Urea Nitrogen

**Figure 1.**
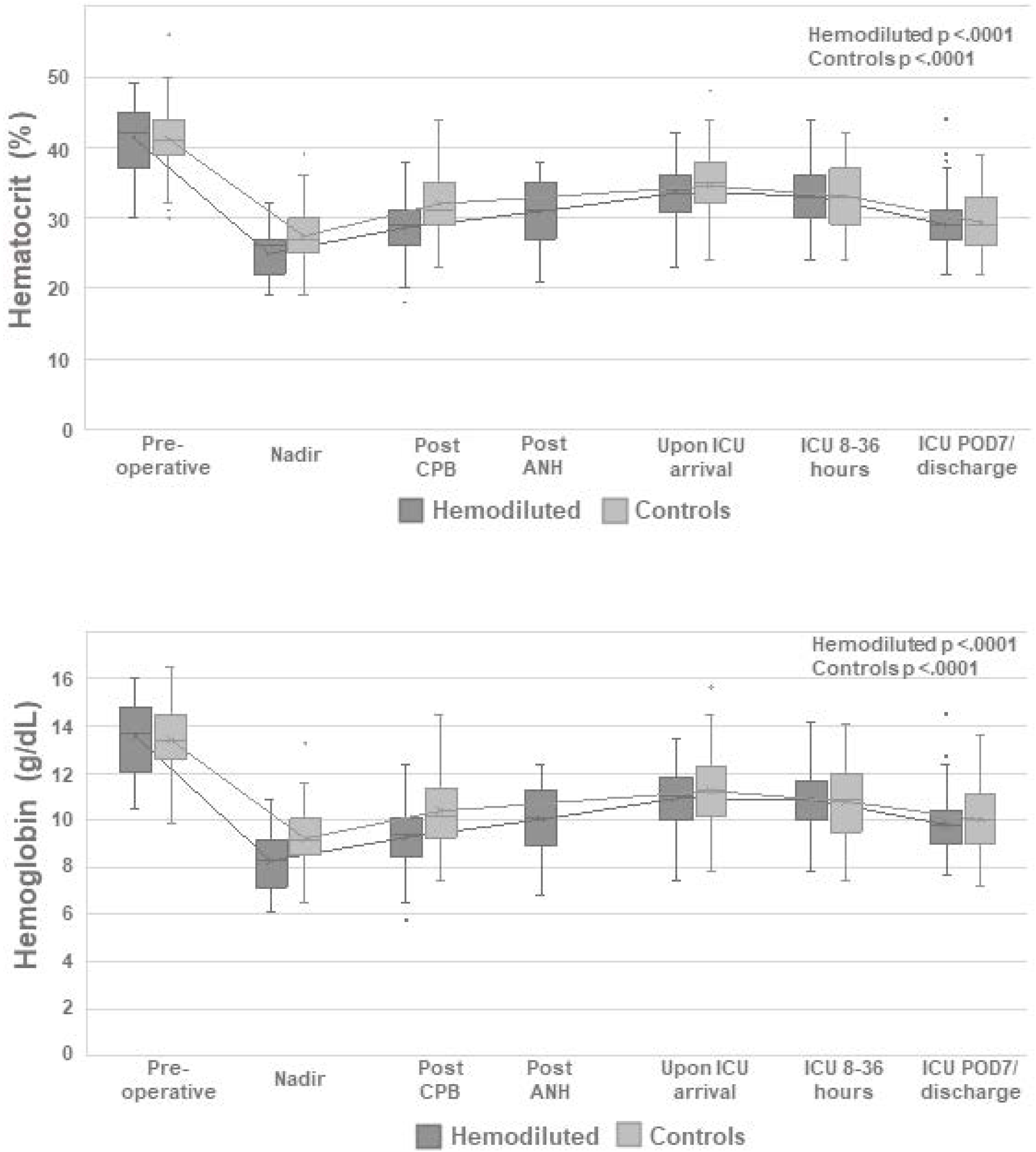
Hematocrit and hemoglobin in the hemodiluted and control groups in the pre-, intra- and post-operative periods. Key: CPB=Cardiopulmonary bypass, ANH=Acute normovelmic hemodilution, ICU=Intensive care unit; *∼50% of the data is missing; Analysis of variance p values compare differences in mean laboratory values across the different time points in each group.

**Figure 2.**
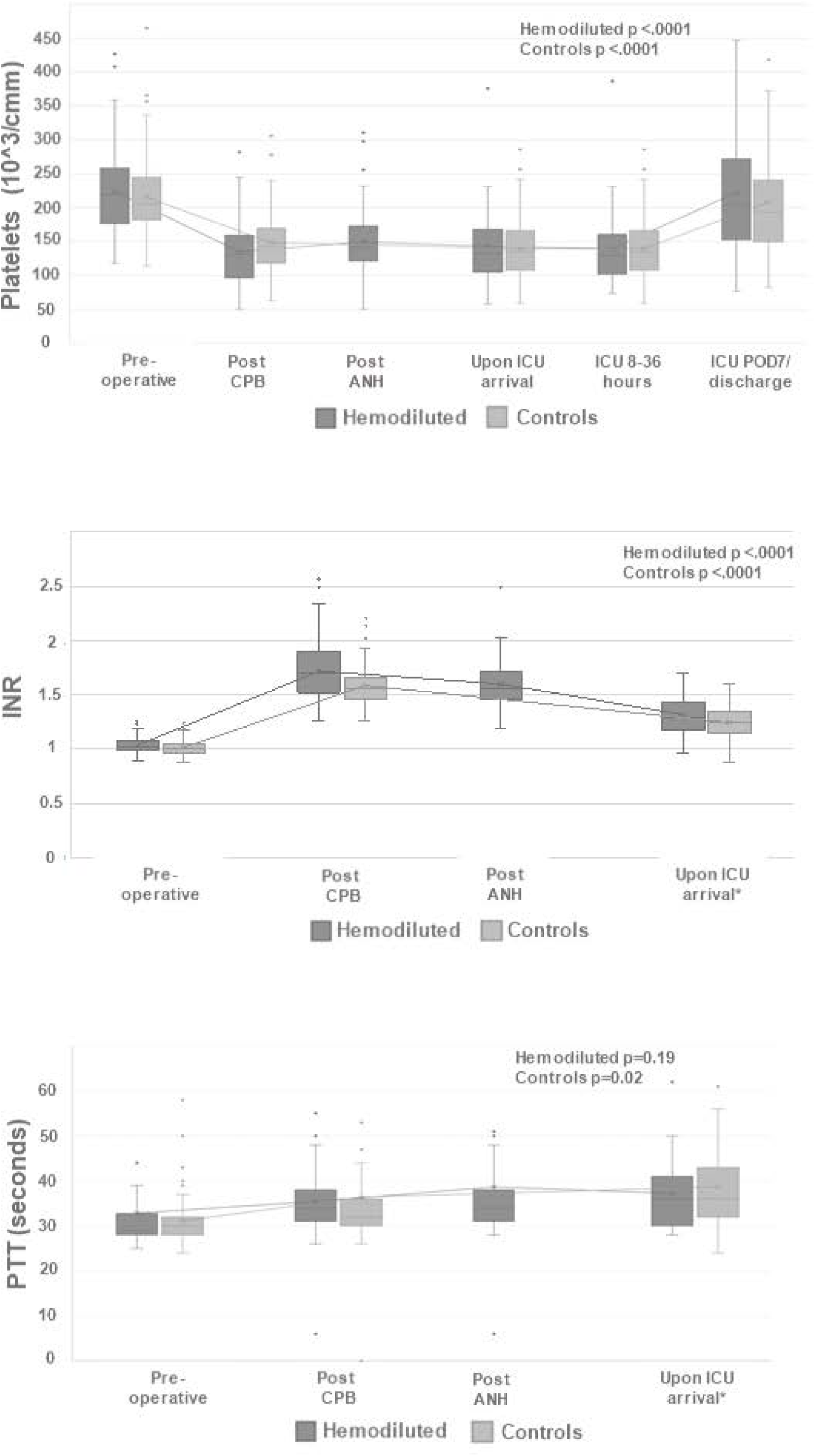
Platelet count, INR and PTT in the hemodiluted and control groups in the pre-, intra- and post-operative periods. Key: INR=international normalized ratio, PTT=partial thromboplastin time, CPB=Cardiopulmonary bypass, ANH=Acute normovelmic hemodilution, ICU=Intensive care unit; *∼50% of the data is missing (ICU arrival); Analysis of variance p values compare differences in mean laboratory values across the different time points in each group.

To assess effects of hemodilution use on hemodynamic interventions, we compared the use of intravenous fluids and vasopressor medication in the ANH and control groups (Table 4). Volume of administered crystalloids was numerically greater in the ANH group, but did not reach statistical significance (*p*=0.22). Volume of intraoperatively administered 25% albumin was significantly higher in the ANH group (116 ± 124 vs. 79 ± 81 ml, *p*=0.04). During the hemodilution period (between removal and return of autologous blood), the ANH group required significantly higher doses of vasopressors compared to control: the highest infusion rate expressed in norepinephrine equivalents was 0.17 ± 0.1 vs. 0.12 ± 0.1 µg/kg/min, *p*=0.01, respectively. Following the period of hemodilution, by the end of the procedure there was no difference in vasopressor requirements: 0.03 ± 0.003 vs. 0.03 ± 0.004 µg/kg/min, *p*=0.95, in ANH vs control, respectively.

**Table 4.** Intra-operative laboratory, physiological parameters and post-operative surgical outcomes for patients who were hemodiluted and controls

|  | Hemodiluted<br>N=66 (39.1%) | Controls<br>N=103 (60.9%) | *p* value |
| --- | --- | --- | --- |
| ***Lactic acid (mmol/L)*** |  |  |  |
| Before CPB | 1.1 ± 0.4 | 1.1 ± 0.4 | 0.24 |
| Peak on CPB | 1.6 ± 0.7 | 1.6 ± 0.9 | 0.96 |
| Last operating room value | 2.2 ± 0.8 | 2.2 ± 1.0 | 0.84 |
| ***Highest Intra operative Right NIRS (%)*** |  |  |  |
| Before hemodilution | 71.7 ± 13.1 | - | - |
| After hemodilution or before CPB | 69.7 ± 13.5 | 73.1 ± 13.3 | 0.11 |
| On CPB | 58.1 ± 13.1 | 62.6 ± 13.8 | 0.03 |
| Last operating room | 65.5 ± 13.4 | 66.1 ± 13.6 | 0.79 |
| ***Highest Intra operative Left NIRS (%)*** |  |  |  |
| Before hemodilution | 72.2 ± 12.5 | - | - |
| After hemodilution or before CPB | 70.4 ± 13.1 | 73.3 ± 11.7 | 0.14 |
| On CPB | 59.1 ± 12.5 | 63.0 ± 13.1 | 0.05 |
| Last operating room | 66.0 ± 13.5 | 67.0 ± 12.2 | 0.62 |
| ***Highest infusion rate of vasopressors in Norepinephrine equivalents (µg/kg/min)*** |  |  |  |
| Before and on CPB | 0.17 ± 0.1 | 0.12 ± 0.1 | 0.01 |
| Last intra-operative | 0.03 ± 0.03 | 0.03 ± 0.04 | 0.95 |
| Total crystalloids given (ml)* | 1739.8 ± 652.8 | 1610.0 ± 542.1 | 0.22 |
| Total albumin given (ml)* | 116.3 ± 124.1 | 78.6 ± 81.0 | 0.04 |
| Urine output in the operating room (ml) | 593.2 ± 307.9 | 514.2 ± 262.1 | 0.07 |
| 12 hour chest tube output (ml) | 380.2 ± 114.1 | 374.9 ± 163.0 | 0.81 |
| Re-operation for bleeding | 0 | 0 | - |
| Mechanical ventilation (hours) | 3.9 [3.2-5.0] | 3.7 [2.9-4.9] | 0.48 |
| ICU length of stay (hours) | 25.5 [21.8-46.6] | 26.0 [21.7-40.8] | 0.29 |
| Acute kidney injury<sup>^</sup> | 15 (22.7) | 29 (28.1) | 0.43 |
| Need for RRT | 0 | 0 | - |
| Infection during this admission | 0 | 1 (0.9) | - |
| Ischemic Stroke | 0 | 1 (0.9) | - |

Key: N (%); medians [Q1-Q3]; mean ± SD; <sup>^</sup>Acute kidney injury was defined as an increase in 0.3 mg/dL or a 1.5 time increase from pre-operative creatinine to post-operative peak creatinine. ICU=Intensive Care Unit, RRT=Renal replacement therapy, CPB=Cardiopulmonary Bypass, NIRS=Near-infrared spectroscopy
\*10-30% of the data is missing

As a surrogate for organ perfusion and oxygenation, lactic acid, urine output and cerebral oximetry (NIRS) were assessed in the two groups (Table 4). No differences in peak lactic acid were detected before CPB, during CPB and at the end of the procedure. There was no difference in intraoperative urine output. During the hemodilution period before initiation of CPB there was no statistical difference in the peak left and right NIRS oxygen saturations. However, during CPB statistically significant lower cerebral oxygen saturations (%) were detected in the ANH compared to control group: right side 58 ± 13 vs. 63 ± 14, *p*=0.03; left side 59 ± 13 vs. 63 ± 13, *p*=0.05, respectively. Following CPB, the difference in cerebral oximetry was no longer statistically significant.

Secondary outcomes including post-operative 12h chest tube output, reoperation for bleeding, duration of intubation, duration of ICU stay, acute kidney injury, need for renal replacement therapy, ischemic stroke and infection rate were not significantly different between the hemodilution and control groups, however some p values were not computed due to small or non-existent events (Table 4).

## DISCUSSION

This is the first study to describe effects of intra-operative hemodilution on physiological parameters in complex cardiac surgeries. It also analyzes laboratory values, as well as major post-operative outcomes. Overall, our results suggest that ANH is not associated with physiologically significant intra-operative reduction in organ perfusion and oxygen delivery. The study also indicates that hemodilution may not be associated with increased incidence of adverse outcomes and produces comparable coagulation laboratory values to other standard practices which advocates its safety as a blood conservation technique for high risk cardiac procedures.

Advancing blood conservation strategies in cardiac surgery remains of great importance as association between blood transfusion and increased morbidity, mortality and healthcare cost has been well established. ^9 10^ Despite being used for decades, benefits of ANH remain controversial, with a supporting level of evidence B and a recommendation class IIb per the Society of Thoracic Surgeons guidelines. ^14 15^ It is important to note that this blood conservation technique has been under-studied in complex and higher risk cardiac surgeries, compared to less complex procedures such as CABG, single valve, or concomitant CABG and single valve procedures. ^3 4^ However, recent studies showed ANH to be associated with reduced transfusions in complex thoracic aortic repairs. ^8 16 17^

Possible reasons for relative under-utilization of hemodilution in complex surgeries are concerns for hemodynamic instability during and after removal of autologous blood, as well as decreased oxygen delivery during the hemodilution period. ^18 19 20^ Hence, rather than studying transfusion-sparing benefits of ANH, we focused on assessing perturbations in laboratory, physiological parameters and outcomes associated with hemodilution in complex cardiac surgeries. By including only hemodiluted and control patients who have not received any allogeneic blood products, we attempted to eliminate the confounding effect that allogeneic blood may have had on intraoperative hemodynamics and post-operative outcomes; and to unmask potentially detrimental consequences of ANH, the study only included patients who had not received any allogeneic blood during their hospitalization. This approach has not been described in the previous literature, and it is considered to be an important step toward understanding of the risk profile of ANH in complex cardiac patients.

Standard coagulation tests obtained immediately after separation from CPB and protamine administration suggest that hemodilution worsens coagulopathy associated with cardiac surgical procedures and CPB. INR was higher and platelet count lower in samples from the ANH group before autologous blood was returned, compared to samples from the same group after blood return or the control group. Those laboratory values equalized between the control and ANH groups after blood return, while fibrinogen remained lower in the ANH group at both time points compared to control. Those findings are consistent with previously published results by others. ^21 22 24^ As fibrinogen in both groups was over 200 mg/dL it is unclear if the statistical difference also translates into significant biological difference, and particularly so in absence of significant clinical bleeding. Importantly, we found no statistically significant difference in the 12 hour chest tube output or rate of re-operation for bleeding in the studied cohorts.

Fluctuations of hemoglobin and hematocrit through the study period demonstrated an expected pattern: pre-operative values being the highest, with nadir during CPB (Figure 1). The values during different pre- and intra-operative time points are consistent with data from the literature. ^22 25^ While some studies demonstrated higher post-operative hemoglobin and hematocrit in ANH compared to control, there was no statistical difference in our cohorts which may be due to study exclusion criteria, as discussed above. ^25^ Importantly, in our study groups hemoglobin and hematocrit differed only during the hemodilution period, which supports the validity of the study design and conclusions that focus on effects of intra-operative hemodilution.

In an attempt to describe and understand how intra-operative hemodilution may be affecting hemodynamics we analyzed the use of intravenous fluid therapy and vasoactive medications at different stages of surgery. Compared to the control, the ANH group received significantly more albumin, while the increase in crystalloids did not reach statistical significance. Increased use of fluids in the ANH group is expected, as it is inherent to the technique of hemodilution. In our institutional practice 25% albumin is commonly used; however it is important to acknowledge that the total amount of fluids, the use of albumin vs. crystalloids, or 5% albumin vs. 25% albumin, are all variable across institutions and individual providers. ^25 26^ Vasopressor medication therapy was assessed by determining the highest infusion rates expressed in norepinephrine equivalents through two time periods: during hemodilution and after hemodilution (at the end of the procedure). The period before hemodilution was not analyzed as only hemodynamically stable patients with no baseline vasoactive medication requirements were included in the study. Significantly higher doses of vasopressors were used in the ANH group during hemodilution compared to controls during the same time-frame period, which may be explained by a relatively inadequate volume expansion (fluid therapy) and/or reduced systemic vascular resistance and blood viscosity associated with hemodilution. ^1^ After the autologous blood was returned, vasopressor requirements between the two groups equalized – before leaving the operating room vasopressor infusion rates were nearly identical.

During hemodilution when intravascular volume and organ perfusion remain adequate, oxygen delivery is also maintained. ^27^ However, even when hemodilution is associated with a decreased oxygen delivery (to a certain degree), the oxygen consumption remains appropriate due to increased oxygen extraction. ^28^ As surrogate measures for adequacy of tissue perfusion and oxygenation, we analyzed regional cerebral oxygen saturation (rSO_2_), urine output, creatinine and lactic acid. NIRS is frequently used in cardiac surgery to provide an estimate of oxygen supply and demand in the frontal region of the head, with described mean baseline between 65% and 68%, which is consistent with our results. ^29 30^ Per our standard practice, rSO_2_ was continuously monitored through the procedure and recorded (charted) every 5 min. As rSO_2_ was not continuously recorded, we opted to analyze the highest values during different stages of the procedure. Analysis of the lowest values would have required elimination of erroneously low readings (due to technical or procedure-related events), a process likely to introduce selection bias. In our study, hemodilution did not result in statistically significant drop in rSO_2_ during the pre-CPB hemodilution period, however during CPB the ANH group had statistically lower rSO_2_. As the difference was modest and limited to the CPB period, it is unclear if such drop in rSO_2_ has clinical significance. No increase in postoperative ischemic stroke was detected, however a more refined neurological and cognitive assessment may need to be done to determine clinical significance. ^31 32^

Acute kidney injury is a common complication of cardiac surgery, that likely results from a mismatch of renal oxygen supply and demand during CPB. ^33 34^ As lower hematocrit reduces oxygen delivery, it is important to analyze the association between hemodilution and kidney injury. There was no difference in post-operative AKI between ANH and control, when AKI was defined as a creatinine increase by 0.3 mg/dL or 1.5 times from the bassline pre-operative value. Also, there was no significant difference in urine output. As our cohorts had a relatively short length of CPB, the sample size may have been underpowered to detect difference in AKI. Increased amount of evidence suggests importance of factors other than hematocrit in providing adequate oxygen delivery. ^34 35^

As with insufficient oxygen supply there is an increased contribution of anaerobic metabolism to the metabolic needs, we studied peak lactic acid before CPB (before hemodilution), during CPB, and at the end of the procedure (after autologous blood return). Values of lactic acid were almost identical in the ANH and control groups, and up-trended in similar fashion through the procedure.

### Limitations

All limitations that are inherent to observational cohort studies apply. Despite stringent patient selection criteria, selection bias remains possible. The small sample size in each group included in the study hindered the ability to conduct analysis that allowed to control for all possible confounders and a possible type 2 error. Inclusion of patients who underwent complex cardiac procedures without receiving allogeneic blood transfusion during their hospitalization limited the number of study subjects. There were no baseline differences within the cohort, however, the increased prevalence in coronary artery disease in the past medical history of the control group may have affected the results. Furthermore, exclusion of all patients with severely reduced ventricular function, and no statistical difference in the proportion of patients who required CABG could suggest that severity of coronary disease was similar between groups. The relatively recent introduction of ANH in our institutional practice allowed for a two-year study period, however it also resulted in a contemporary and consistent clinical practice through the duration of the study. Although authors believe that point of care coagulation tests (TEG^®^) do have a role in assessing coagulopathy, those tests have not been consistently used in our institutional practice and we were unable to include them in the analysis due to the level of missing data. ^22 23 24^ Interpretation of coagulopathy and laboratory values in cardiac surgery remains challenging, and particularly so in our study group, as one of the frequently used indicators of coagulopathy is number of transfused blood products. Secondly, by excluding patients who received allogeneic transfusion, we likely selected for patients with milder coagulopathy and uncomplicated procedures (i.e., shorter CPB runs).

### Conclusion

This study demonstrates that in our practice ANH is associated with increased intra-operative administration of intravenous fluids as well as vasopressor medications during the hemodilution period. It also suggests that this blood conservation technique does not have significant adverse effects on intra-operative organ perfusion or oxygen delivery, as well as major post-operative outcomes. Results indicate that ANH can be safely applied in complex procedures, and concerns for hemodynamic instability and impaired tissue oxygenation during hemodilution should not hinder its utilization in complex cardiac surgeries. Overall results provide information that may be important in identifying patients in which harmful effects of hemodilution may outweigh its benefits. Those insights would be of great value in further refinement and wider implementation of ANH protocols in complex cardiac surgeries.

## Data Availability

The data that support the findings of this study are available from the corresponding author (D.M.) upon reasonable request.

## ACKNOWLEGDEMENTS

Informatics services for this project were provided by the Anesthesiology Informatics group, a resource supported by the Department of Anesthesiology and Perioperative Medicine, University of Alabama at Birmingham.

